# Selection bias as an explanation for the observed protective association of childhood adiposity with breast cancer

**DOI:** 10.1101/2022.12.08.22283258

**Authors:** C M Schooling, K Fei, J V Zhao

## Abstract

**Objective:** Recalled childhood adiposity is inversely associated with breast cancer observationally, including in Mendelian randomization (MR) studies, questioning its role. Breast cancer studies recruited in adulthood only include survivors of childhood adiposity and breast cancer. We assessed recalled childhood adiposity on participant reported sibling and maternal breast cancer to ensure ascertainment of non-survivors using MR.

**Study Design and Setting:** We obtained independent strong genetic predictors of recalled childhood adiposity for women and their associations with participant reported own, sibling and maternal breast cancer from UK Biobank genome wide association studies (GWAS). We obtained MR inverse variance weighting estimates.

**Results:** Childhood adiposity in women was inversely associated with own breast cancer (odds ratio (OR) 0.66, 95% confidence interval (CI) 0.52 to 0.84) but unrelated to participant reported sibling (OR 0.85, 95% CI 0.60 to 1.20) or maternal breast cancer (OR 0.84, 95% CI 0.67 to 1.05 respectively).

**Conclusion:** Weaker inverse associations of recalled childhood adiposity with breast cancer with more comprehensive ascertainment of cases before recruitment suggests the inverse association of recalled childhood adiposity with breast cancer is due to selection bias arising from preferential selection of survivors. Greater consideration of left truncation in public health relevant causal inferences is warranted.

**Highlights:** Recalled childhood adiposity is inversely associated with breast cancer.

Studies of childhood exposures recruited in adulthood are open to left truncation.

Participant reports about family members include deaths before recruitment.

We tested childhood adiposity on sibling breast cancer using Mendelian randomization.

Childhood adiposity with sibling breast cancer was null, suggesting left truncation.

## Introduction

Over many years, observational studies have reported recalled childhood adiposity inversely associated with breast cancer,^1-4^ raising questions about the role of adiposity in childhood for girls. Observational studies are open to confounding. As such, these somewhat counter-intuitive findings could be the result of uncontrolled confounding, given the difficulty of enumerating and measuring all potential confounders comprehensively. Recently, a Mendelian randomization (MR) study, which is less open to bias from confounding because it uses genetic proxies of exposure,^5^ also found recalled childhood adiposity inversely associated with breast cancer.^6^ Several plausible reasons have been suggested for childhood adiposity protecting against breast cancer. Greater childhood adiposity could slow pubertal growth and sexual maturation, which could be protective.^4^ Childhood adiposity could also result in a hormonal milieu protective against breast cancer.^4^ Alternatively, the effect of exposure to adiposity during childhood may not be relevant into adulthood,^6^ or may be different, perhaps because the effects of early life experiences are only fully actuated after puberty. A very comprehensive MR study, also using recalled childhood adiposity (at ∼57 years), recently reported no biological explanation for the observed inverse association of childhood adiposity with breast cancer,^7^ leaving the inverse association of childhood adiposity with breast cancer an unresolved paradox. Here, we considered another possible explanation: that the observed inverse association of childhood adiposity with breast cancer is bias rather than a target of intervention.

Observational studies, apart from being open to confounding, are also open to selection bias,^8^ particularly for exposures, such as childhood adiposity, that may occur well before recruitment.^9^ Specifically, a study of the effect of childhood adiposity on breast cancer recruited in adulthood is missing the women who died from childhood adiposity before recruitment and the women who died of breast cancer before recruitment, particularly given breast cancer causes death relatively early in adulthood.^10^ Only studying those who have survived to recruitment, possibly years after exposure, creates a specific form of selection bias, i.e. survival bias, or depletion of susceptibles,^11^ because of incomplete ascertainment particularly of exposed cases. Currently, few methods of addressing survival bias exist, although sensitivity analysis is possible.^12, 13^

Here, we used an MR study to assess the association of childhood adiposity with participant reported sibling and maternal breast cancer to obtain more comprehensive ascertainment of deaths from breast cancer before recruitment, given an MR study reduces bias from confounding.^5^ Using a design based on participant reports of sibling or maternal breast cancer reduces survival bias by including breast cancer cases even if the case died before recruitment of the participant (Figure 1b compared with Figure 1a). So, associations of childhood adiposity with participant reported sibling or maternal breast cancer should give a less biased estimate for childhood adiposity on breast cancer. We also assessed the potential relevance of survival bias due to death before recruitment from childhood adiposity or breast cancer, from the associations of childhood adiposity and liability to breast cancer with lifespan. The weaker the association for liability to breast cancer the more likely the GWAS is missing deaths before recruitment given breast cancer is a major cause of premature mortality in women.^14^

**Figure 1:**
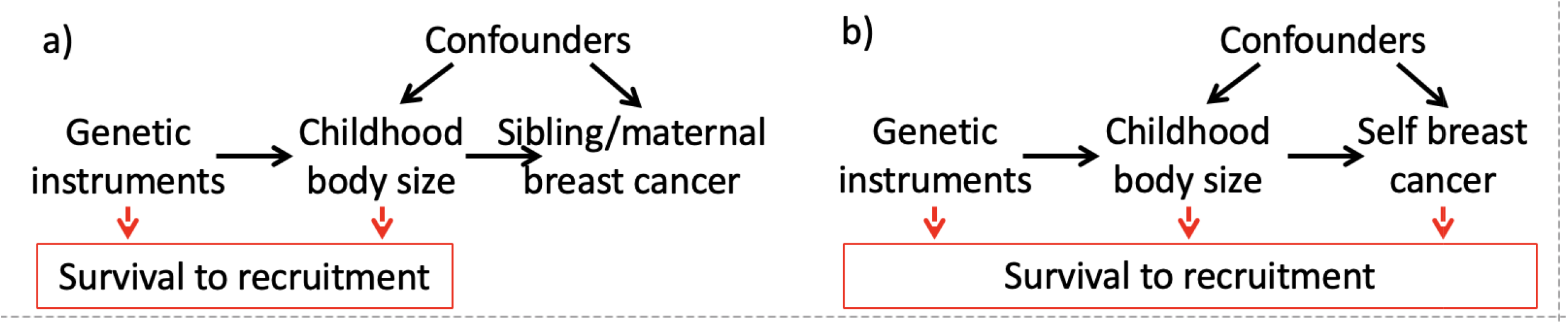
Directed acyclic graph showing that a Mendelian randomization study of childhood adiposity on self breast cancer recruited in middle to older age is biased by selection on genetic endowment and surviving breast cancer (Figure 1b) but a study of participant reported sibling or maternal breast cancer is likely less biased because it does not select on surviving breast cancer to recruitment (Figure 1a). A box indicates selection on an attribute.

## Materials and Methods

MR as a form of instrumental variable analysis requires the assumptions of relevance, independence, and exclusion restriction. To address relevance, we only selected as instruments genome wide significant (p-value<5×10^−8^) independent (r^2^<0.001) genetic variants for each exposure. Use of genetic instruments helps ensure independence from confounders.^15^ The exclusion restriction assumption is more difficult to satisfy because it can be violated by genetic pleiotropy or selection bias, including survival bias.^13^ We assessed genetic pleiotropy by sensitivity analysis. We addressed potential selection bias for childhood adiposity on breast cancer by using participant reported sibling and maternal breast cancer as outcomes, and from the relevance of potentially biasing pathways (Figure 1), i.e., childhood adiposity and breast cancer on survival to recruitment proxied by lifespan. We used two-sample MR, where possible. Some questions could only be addressed using one-sample MR which can be biased even for very large samples.^16^ So, we assessed the validity of one-sample MR estimates from sample size and relevant diagnostics, such as I^2^_GX_.^16^

### Study design

This is mainly a one-sample MR study using genome wide association studies (GWAS) of the UK Biobank. The UK Biobank is a population-based cohort of half a million people, intended to be recruited at age 40 to 69 years, average age ∼57 years, from 22 sites in Great Britain in 2006 to 2010.^17^ The participants completed extensive questionnaires, as well as undergoing tests and providing samples for genotyping and other investigations. The UK Biobank includes information on childhood adiposity (relative body size at 10 years), breast cancer in self and family members (siblings and mothers), as well as sex-specific parental survival (age at death or current age). Quality controlled GWAS for up to ∼13.5 genetic variants are publicly available, both sex-combined^18^ and sex-specific^19^ (http://www.nealelab.is/uk-biobank). We also used the largest publicly available GWAS of self breast cancer from the Breast Cancer Association Consortium (BCAC),^20^ as used previously,^21^ which is largely based on case-control studies from Europe and North America.

### Data sources

#### Childhood adiposity

We obtained genetic instruments for childhood adiposity from a UK Biobank GWAS of self-reported comparative childhood body size at age 10 years (thinner, about average, plumper) from the UK Biobank,^17^ concerning 191344 white British women adjusted for principal components, age and age^2^ (http://www.nealelab.is/uk-biobank).

#### Breast cancer

We used a quality controlled GWAS of participant reported sibling breast cancer (cases=16586, non-cases=345233) and mothers breast cancer (cases=35102, non-cases=388356) of white Europeans from the UK Biobank,^18^ obtained using linear regression after adjusting for principal components. These estimates were converted from probability into log odds using an approximation.^22^ Given siblings and mother only share half their genetic endowment with the participant, these estimates were also doubled. For comparison, we also used a similarly obtained GWAS of self breast cancer from the UK Biobank (cases=10303, non-cases=452630) and the BCAC GWAS of self breast cancer (cases=122977, controls=105974) largely in Europeans which was quality controlled and adjusted for country and principal components.^20^

#### Lifespan

We used a GWAS of mother’s attained age from the UK Biobank for lifespan,^19^ based on reports of mother’s attained age (age at death or current age), which is partly heritable.^23^ Only white Europeans were included.^24^ Deaths of mothers before 57 years and adopted participants were excluded.^24^ Quality controlled genetic associations with Martingale residuals from Cox proportional hazards were adjusted for age, assessment centre and array type.^24^ Estimates were converted into life years using an established approximation.^25^

### Statistical analysis

#### Genetic instruments

We selected genome wide significant (p-value <5×10^−8^) independent (r^2^<0.001) genetic variants as instruments for all exposures considered. As a measure of instrument strength, we obtained each F-statistic as the estimate for genetic variant on exposure divided by its variance.^26^ We aligned genetic variants across studies on the same allele letter, using effect allele frequency as necessary after dropping palindromic variants with minor allele frequency >0.42.

#### Analysis

We used inverse variance weighting (IVW) as the main analysis. We used the weighted median (WM) and MR-Egger (MRE) as sensitivity analysis to assess potential violation of the exclusion restriction assumption by pleiotropy or selection bias. The WM gives a valid estimate as long as at least 50% of the weight is based on valid instruments.^27^ MRE gives valid estimate when the genetic instruments do not affect confounders of exposure on outcome,^26^ but has low power.

MR-Base was used to extract instruments and align genetic variants across studies,^18^ the MendelianRandomization R package was used to obtain estimates. All analyses was conducted using R (version 4.1.2).^28^ This study only uses genetic summary statistics created from information and materials previously collected with informed consent.

## Results

At least 75 genetic variants with F-statistics greater than 10 and average F-statistic >66 were obtained for childhood adiposity in women and were available for all outcomes (Table 1). Childhood adiposity was not clearly related to UK Biobank participant reported sibling breast cancer or to UK Biobank participant reported mother’s breast cancer. In contrast, childhood adiposity was strongly inversely associated with self breast cancer from UK Biobank and BCAC (Table 1).^20^ Sensitivity analysis, i.e., the MR Egger intercept, indicated that the estimates for childhood adiposity on breast cancer from BCAC might be invalid (Table 3) due to pleiotropy or selection bias.

**Table 1:**
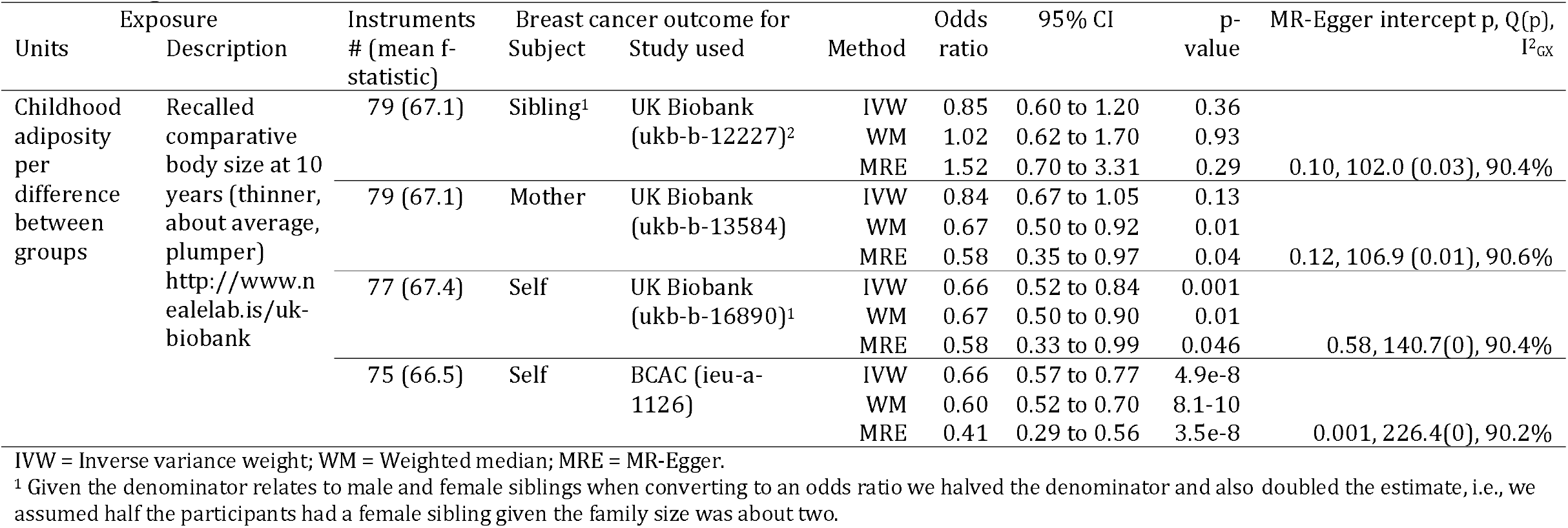
MR estimates in women for childhood adiposity from the UK Biobank on breast cancer from UK Biobank participant reports about siblings and mothers and on self breast cancer from the UK Biobank and BCAC.

**Table 2:** MR estimates in women for childhood adiposity on lifespan (life years) in the UK Biobank.

| Units | Subject | Exposure |  | n | Method | Instrument # (mean f-statistic) | Outcome – Lifespan (ebi-a-GCST006696) |  |  | MR-Egger intercept p-value, Q(p), I<sup>2</sup><sub>GX</sub> |
| --- | --- | --- | --- | --- | --- | --- | --- | --- | --- | --- |
|  |  | Description |  |  |  |  | Life years<sup>1</sup> | 95% CI | p-value |  |
| Childhood adiposity per difference between groups<sup>2</sup> | Self | Comparative body size at 10 years |  | 191344 | IVW | 79 (67.1) | -1.67 | -2.71 to -0.64 | 0.002 | 0.22, 174 (0), 90.3% |
|  |  | http://www.nealelab.is/uk-biobank |  |  | WM |  | -1.25 | -2.44 to -0.06 | 0.04 |  |
|  |  | biobank |  |  | MRE |  | -2.98 | -5.31 to -0.65 | 0.01 |  |

IVW = Inverse variance weight; WM = Weighted median; MRE = MR-Egger.
<sup>1</sup> Years of life obtained from the log protection ratio multiplied by 2.5863 to account for children only sharing half their genetic endowment with their mothers and then by 10 as a verified actuarial rule of thumb <sup>19</sup>
<sup>2</sup>Thinner, about average, plumper compared to others at age 10 years

**Table 3:**
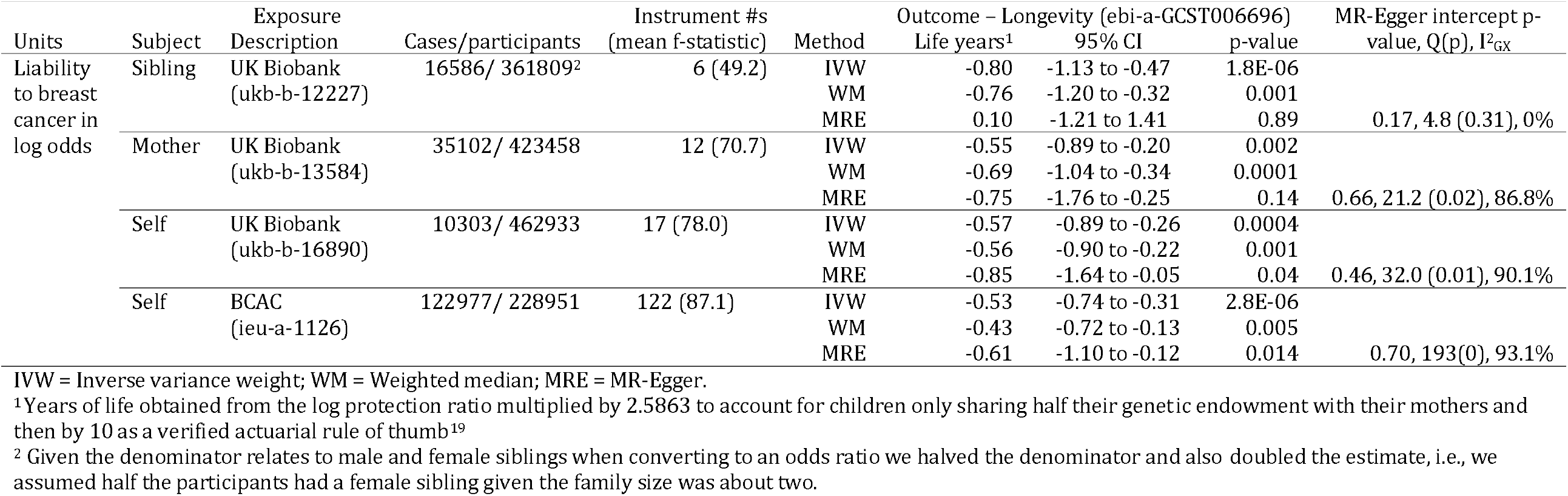
MR estimates in women for liability to breast cancer from UK Biobank participant reports about siblings and mothers and for self from the UK Biobank and BCAC on lifespan (life years)

As expected, childhood adiposity in women was strongly inversely associated with lifespan in women (Table 2). MR estimates were similar from all sensitivity methods, and the MR-Egger intercept p-value did not indicate pleiotropy (Table 2).

Associations of liability to breast cancer with lifespan differed by breast cancer study (Table 3), with the greatest magnitude of association for participant reported sibling breast cancer from the UK Biobank and the smallest magnitude of association for BCAC (Table 3). The F-statistics were adequate. Sensitivity analysis gave a similar interpretation, with no evidence of pleiotropy from the MRE intercepts (Table 3).

## Discussion

Childhood body size was not clearly associated with breast cancer using participant report of siblings’ or mothers’ breast cancer to obtain more complete ascertainment of lifetime breast cancer. Moreover, we showed the existence of biasing pathways when considering effects of a harmful early life exposure on a disease that can cause death relatively early in adulthood. Collectively, this study shows that the inverse association of childhood adiposity with breast cancer could be an artefact of selection bias arising from only selecting survivors. Moreover, the finding of no association of childhood adiposity with participant reported sibling’s breast cancer or mother’s breast cancer is consistent with a recent study which found no physiological reason why childhood adiposity should protect against breast cancer,^7^ but, parsimoniously, suggests the explanation is a bias.

Previous studies have emphasized the importance, when assessing potentially causal effects, of ensuring the exposure starts at or after recruitment to avoid biased estimates,^9^ as well as the importance of counting exposure time correctly for the exposed and unexposed.^29^ The issues raised by some potential participants being missing from a study because of prior death due to the exposure or outcome, or a competing risk of the outcome^30^ is known,^31-33^ sometimes as left truncation, and is often thought to have most relevance for studies in the elderly.^31-33^ In contrast, population representative studies are widely considered to be a keystone of scientific research.^34^ So bias in associations for long-term harmful exposures in studies only representing survivors of the original birth cohorts who formed the population may seem counter-intuitive.

Despite using MR to reduce confounding, using participant reported sibling and maternal breast cancer to address selection bias and conducting additional analysis to check the conditions for selection bias, this study has limitations. First, childhood adiposity was self-reported in adulthood in three groups, so may not be entirely accurate, possibly reducing precision which is compensated for by the large sample size. Second, lifespan was based on participant (adult child) reported maternal attained age which could be open to measurement error, potentially biasing towards the null. However, reports about others can be more reliable than self-reports^35^ and mother’s age is also important. Mothers who died early (before age 57 years) were excluded, likely biasing towards the null or the reverse of the true association. Third, sibling’s and mother’s breast cancer could be reported imprecisely, which would bias estimates towards the null. Mother’s breast cancer could also be open to survival bias due to competing risk, where in older women death from cardiovascular disease might preclude the occurrence of breast cancer. Nevertheless, liability to sibling breast cancer had the largest association with life years lost, as expected. Fourth, publicly available GWAS of sibling breast cancer by sex (of the sibling) are not available, making the magnitude of the estimates for sibling breast cancer subject to several assumptions. Fifth, the UK Biobank is not population representative, which would be a source of bias for a descriptive study but is less likely to be a source of bias for MR studies unless the participants were selected on genetic endowment and the outcomes. The BCAC GWAS of breast cancer is a meta-analysis of case series, case-control studies and cohort studies carefully designed to avoid genetic confounding, but less explicitly designed to address selection bias.^20^ Sixth, MR studies rest on the assumptions of relevance, independence and exclusion. The genetic instruments strongly predicted each exposure, with high F-statistics. Random allocation of genetic material at conception reduces the risk of the genetic instruments being associated with confounders. However, there remains a possibility of violating the exclusion restriction assumption by selecting on childhood BMI. Such bias is not thought to be very influential,^36^ but would bias towards a protective effect of childhood BMI. Seventh, one-sample MR, which can be biased towards the confounded estimate, was largely used. However, differences were found between estimates for childhood adiposity on breast cancer from participant self-reports and participant reports about their family (siblings and mother) in the UK Biobank when confounding is likely to be similar for self, siblings and mother. Moreover, the large sample sizes suggest any such bias is likely to be minimal for IVW and WM estimates.^16^

Here, we demonstrate that when using observational studies to examine the effect of exposures that start before recruitment it is important to consider whether selection bias due to differential survival might have occurred because of inadvertent exclusion of deaths before recruitment. Use of MR studies with suitable proxy outcomes,^37^ such as siblings or parents, may help identify bias arising from inadvertently focusing on survivors. However, in this context it is vital to obtain information about all siblings/mothers regardless of mortality status rather than only including or recruiting those who survived to recruitment, which would not address selection bias due to only selecting survivors. Overall, our study draws attention to the importance of considering whether studies recruited in adulthood, particularly of lifetime exposures, such as genetics, could be biased by almost inevitable selection on survival. Bias is likely to be most marked when selecting on survival of exposure and outcome,^38^ or on survival of exposure and a competing risk of the outcome.^13, 30^ Survival bias is particularly relevant for studies, such as genetic studies, where the exposure often commences well before recruitment and the outcome is a disease state, where missing cases may considerably distort the comparisons. In contrast, for continuous outcomes missing a few people at the extremes of the distribution is unlikely to affect comparisons of central tendency.

## Conclusion

Observations of childhood adiposity associated with a lower risk of breast cancer could be an artefact of selection bias. Assessing the risk of selection bias, due to inadvertently only including survivors, by using controls, such as participant reports on family members, and sensitivity analysis could ensure resources are focused on valid questions, and doubt is not cast on important targets of intervention, such as childhood adiposity.

## Data Availability

https://www.mrbase.org
http://www.nealelab.is/uk-biobank

## Ethics Approval

This study only uses publicly available summary statistics, so does not require ethical approval.

## Author contributions

CMS designed the study in consultation with KF. CMS conducted the initial analysis. JVZ and KF reviewed and checked the analysis and results. CMS wrote the initial draft, which JVZ and KF reviewed thoroughly for important intellectual content. All authors gave final approval to this version and are accountable for all aspects of the work.

## Data Statement

The data used here is available in MR-Base, apart from the GWAS of childhood adiposity in women which is available from Neale Lab http://www.nealelab.is/uk-biobank

## Acknowledgements

None

## Author Contributions

CMS conceptualized the study and the methodology, conducted the initial analysis and wrote the original draft. All authors discussed and reviewed the study methodology in detail. KF and JVZ checked the analysis, edited and reviewed the final version critically for important intellectual content and approved the final version for submission.

Replication can be achieved using MR Base https://www.mrbase.org, publicly available data and R code available on request

